# Differential profile of Leptospirosis in patients under and over 5 years of age in health centers of the Peruvian Amazon, 2023–2024

**DOI:** 10.1101/2025.08.14.25333657

**Authors:** Stefano V. Davila-Philipps, Kary K. Vela-Tello, Jorge I. Carrasco-Celi, Yoki N. Rios-Alava, James L. Vasquez-Lechuga, Marcos H. Parimango-Alvarez, Edgar A. Ramirez-García, Johan Marin-Lizarraga, Tery Vasquez Hassinger, Karine Zevallos

**Affiliations:** Universidad Nacional de la Amazonía Peruana, Iquitos, Loreto, Perú; Hospital Regional de Loreto, Iquitos, Loreto, Perú; Facultad de Medicina, Universidad Nacional de la Amazonia Peruana, Iquitos, Loreto, Perú

**Keywords:** Leptospirosis, Children, Signs and Symptoms, Epidemiology, Amazon, Peru

## Abstract

**Background:** Leptospirosis is an emerging zoonotic disease with high incidence in tropical regions such as the Peruvian Amazon. Although it affects individuals of all ages, knowledge about its clinical and epidemiological behavior in children under five years old remains limited. This population may present atypical clinical forms, nonspecific symptoms, and a milder course, increasing the risk of underdiagnosis and delayed treatment.

**Methodology:** A descriptive and retrospective study was conducted by reviewing medical records and epidemiological forms of all MAT-confirmed patients from 2022 to 2024 in primary health centers in Belén, the district with the highest historical prevalence in Loreto. A total of 400 cases out of 1666 reported were included. Among children under five (n=28), clinical presentation was predominantly nonspecific, with malaise (78.6%) and fever (71.4%) being most frequent. Nearly half (46%) sought care within the first three days of symptoms, and 50% lived in flood-prone areas. Nutritional status showed a significant association, with a predominance of underweight (p<0.001), remaining significant in multivariate analysis (OR=0.051; 95%CI: 0.011–0.233; p<0.001). Additionally, each additional day of illness was associated with an increased likelihood of belonging to this age group (OR=1.366; 95%CI: 1.135–1.644; p=0.001). Regarding serovars, *Panama* showed significant association with children under five (p=0.024), whereas *Hurstbridge* was absent.

**Conclusion:** Children under five with leptospirosis exhibit a distinctive clinical and epidemiological profile, characterized by nonspecific symptoms, higher frequency of underweight, and earlier healthcare-seeking behavior. Associations with malaise, poor nutritional status, and specific serovars like *Panama* suggest a less typical presentation, potentially hindering diagnosis at the primary care level. These findings highlight the need for tailored surveillance, diagnostic, and management strategies for this age group, particularly in endemic regions such as the Peruvian Amazon.

**Author summary:** Leptospirosis is a disease transmitted through contact with water or soil contaminated by animal urine, especially in tropical regions like the Peruvian Amazon. While it can affect people of any age, little is known about how it presents in children under five. In our study, we analyzed 400 confirmed cases from health centers in Belén, one of the most vulnerable districts in the region. We found that children under five often had vague symptoms like general discomfort and fever, which are easy to miss or confuse with other common infections. These children were also more likely to be underweight and live in areas affected by seasonal flooding. Unlike older patients, they tended to visit health centers earlier after symptom onset, possibly because caregivers noticed changes quickly. We also found that a specific type of bacteria (the Panama serovar) was more common in this age group. These findings suggest that leptospirosis may behave differently in young children, which can make diagnosis and treatment more challenging. Recognizing these differences can help improve detection and care for young children in endemic regions.

## Introduction

Leptospirosis is a globally distributed zoonotic disease caused by spirochete bacteria of the genus *Leptospira*. It presents a wide clinical spectrum, ranging from subclinical infections to severe cases with multiorgan failure. Transmission to humans occurs through direct or indirect contact with the urine of wild or domestic animals, which serve as reservoirs and sources of infection. Among the most affected species, rodents are considered chronic carriers, along with domestic animals such as dogs, pigs, and cattle. *Leptospira* colonizes the proximal renal tubules of the reservoirs and is excreted in the urine, turning these species into transient or permanent urinary carriers. In our setting, ecological and sanitary factors contribute to the persistence of the bacteria in contaminated soil and water, increasing human exposure, especially in areas affected by flooding or with precarious living conditions [1].

This disease has a worldwide distribution, with the highest incidence in tropical and subtropical countries. Regions with the greatest estimated morbidity and mortality include Sub-Saharan Africa, Latin America and the Caribbean, South and Southeast Asia, and Oceania. The estimated global annual incidence exceeds one million cases, including approximately 59,000 deaths [2].

In Peru, leptospirosis exhibits an endemic-epidemic pattern, with case surges following rainfall and flooding, increasing the risk of outbreaks. Examples include the outbreaks in Loreto during 2012–2013 due to the overflow of major rivers (Itaya, Nanay, and Amazon) [3], as well as those in 2017–2018, triggered by the El Niño phenomenon, and more recently in 2023 due to Cyclone Yaku, mainly affecting the northern coast of the country [4].

As of Epidemiological Week (EW) 52 of 2024, Peru had reported 9,520 cases of leptospirosis, a 33.7% increase compared to 2023. The incidence rate was 27.91 per 100,000 inhabitants, with 16 confirmed deaths, 4 of which were from the Loreto department. Additionally, 41.25% of reported cases were classified as probable. Of all cases reported by EW 52 of 2024, 75% were concentrated in the departments of Loreto, Ayacucho, Madre de Dios, and Ucayali [5].

Within the epidemiological profile of leptospirosis cases, age has been shown to be a variable associated with important clinical and prognostic differences. Although leptospirosis can occur at any age, there is evidence suggesting that clinical presentation and outcomes may vary significantly between children and adults. The clinical manifestations are diverse and resemble those of other febrile illnesses, including tropical diseases such as dengue, rickettsial infections, malaria, and bacterial sepsis [6]. The disease can range from an asymptomatic form to an acute self-limiting febrile illness, or even a life-threatening condition involving multiorgan dysfunction [7]. Most cases present as uncomplicated fever, but up to 10% may progress to severe forms [8]. Some studies suggest that pediatric patients may experience a less severe course compared to adults, often presenting with atypical clinical forms and lower frequencies of jaundice, renal failure, and need for dialysis upon hospital admission [9]. Furthermore, lower mortality rates have been observed in this group, with case fatality rates up to five times lower than in hospitalized adults with severe disease [10]. This difference may be due to multiple factors, including immunological differences, lower burden of comorbidities in children, milder initial clinical presentation, or earlier healthcare-seeking behavior by caregivers [11,10,12]. However, the less typical clinical presentation in children can also delay diagnosis if clinical vigilance is not maintained.

Although there are publications on leptospirosis in general populations, there remains a limited number of studies that systematically compare clinical and epidemiological differences between specific age groups, particularly in children under five years of age. Therefore, the objective of this study is to describe the clinical manifestations and epidemiological characteristics of confirmed leptospirosis cases in patients under and over 5 years of age treated at four primary health care centers in the district of Belén, Loreto region. This differential approach aims to provide useful evidence to strengthen strategies at three levels—prevention, diagnosis, and treatment—with an emphasis on the pediatric population under 5 years old, a group that remains underexplored in endemic contexts such as the Peruvian Amazon.

## Methods

### Study design

A multicenter, descriptive, and retrospective study was conducted. The study population included patients with a confirmed diagnosis of leptospirosis by the Microscopic Agglutination Test (MAT), who were treated at four Level I health centers: Centro de Salud (C.S) 06 de Octubre, C.S Cardozo, C.S 09 de Octubre y C.S Belén in the district of Belén, province of Maynas, department of Loreto, from January 1, 2022, to December 31, 2024.

Patients were stratified by age (categorized as under 5 years and over 5 years), sex, district of origin, type of area (rural, urban, or peri-urban), and residence in flood-prone zones (areas that can be flooded due to rainfall and/or rising nearby rivers depending on the season). Clinical data included signs and symptoms at admission, duration of illness (from symptom onset to presentation at the health center), type of onset (insidious or abrupt), disease course (stationary or progressive), discharge status (alive or deceased), and treatment received (antibiotics, antipyretics, intravenous hydration, and others).

The confirmatory diagnosis of leptospirosis was based on the MAT test specific for *Leptospira*, processed at the National Institute of Health laboratory in Lima. A case was considered confirmed when a single blood sample showed a serological titer equal to or greater than 1:800, also identifying the serological variant.

The district of Belén, located in the province of Maynas, Loreto region, is one of the most vulnerable urban areas of the city of Iquitos. It has a population of approximately 76,000 inhabitants, of whom 75% live in flood-prone areas, particularly in the lower zone of Belén, which borders the Itaya River and surrounding communities. The climate is warm, with temperatures ranging from 17°C to 38°C, humid and rainy, with an annual average rainfall exceeding 2,600 mm [13]. During seasonal river flooding, many homes become partially submerged, leading to unsanitary conditions due to water stagnation and inadequate waste management. These conditions favor the proliferation of vectors and the transmission of infectious diseases, especially among children under 5 years old, who face high rates of acute respiratory infections, intestinal infections, anemia, and parasitic diseases. In this context, leptospirosis emerges as a reemerging disease closely linked to exposure to stagnant water contaminated with rodent urine, in an environment with limited access to potable water services (50.4%). These conditions make Belén a priority area for integrated public health and environmental management interventions.

### Procedure and data collection

Data was collected from medical records of patients with confirmed leptospirosis and the Clinical-Epidemiological Research Form for Leptospirosis from the Technical Health Standard for the Comprehensive Care of People Affected by Leptospirosis. Patients were included in the study if their medical records were adequately completed and the diagnosis of leptospirosis was confirmed by the MAT test.

### Statistical analysis

The collected information was entered into spreadsheets using the Microsoft Excel software program (Windows 10 version). The analysis was performed using the statistical software IBM SPSS Statistics version 27.0, for Windows 11. Descriptive statistics were used to summarize sociodemographic, clinical, and epidemiological variables. Categorical variables were expressed as absolute and relative frequencies (n, %). All clinical and epidemiological variables, including symptom duration, were treated as categorical variables. Bivariate analyses were conducted using Pearson’s chi-square test to identify associations between categorical variables and age groups (<5 vs. ≥5 years). Variables with p-values <0.05 in the bivariate analysis were included in a multivariate model. A binary logistic regression analysis was performed to identify independent factors associated with being under 5 years of age, with results expressed as odds ratios (OR) and 95% confidence intervals (CI). A p-value of <0.05 was considered statistically significant.

### Ethics statement

The study was approved by the Ethics Committee of the Regional Hospital of Loreto “Felipe Santiago Arriola Iglesias” (No. 091-CIE-HRL-2024). Ethical principles such as confidentiality and the exclusive use of anonymized data were guaranteed. The request for informed consent was not considered necessary for this study because the information was collected from clinical records.

## Results

There were 81 cases (20.25%) in 2022, 105 cases (26.25%) in 2023, and 214 cases (53.5%) in 2024 (Table 1). A progressive increase in cases was observed over the study years; notably, in 2024, the number of cases increased by 103.8% compared to 2023. The majority of cases in children under 5 years of age occurred in 2024 (71%), while the proportion in individuals over 5 years old was 52% in that same year. The health center in Belén that reported the highest number of cases during the study period was the C.S 06 de Octubre, with 288 cases (72%), while the health center that reported the fewest cases was the C.S Belén, with 4 cases (1%) (Figure 1) (Table 1). Regarding the monthly distribution, we observed a marked seasonal pattern in patients over 5 years of age, with significant peaks in the months of March (62 cases), September (49 cases), and October (49 cases). In contrast, cases in children under 5 years of age were scarce and dispersed throughout the year, with a maximum of 5 cases in August (Figure 2).

**Table 1.**
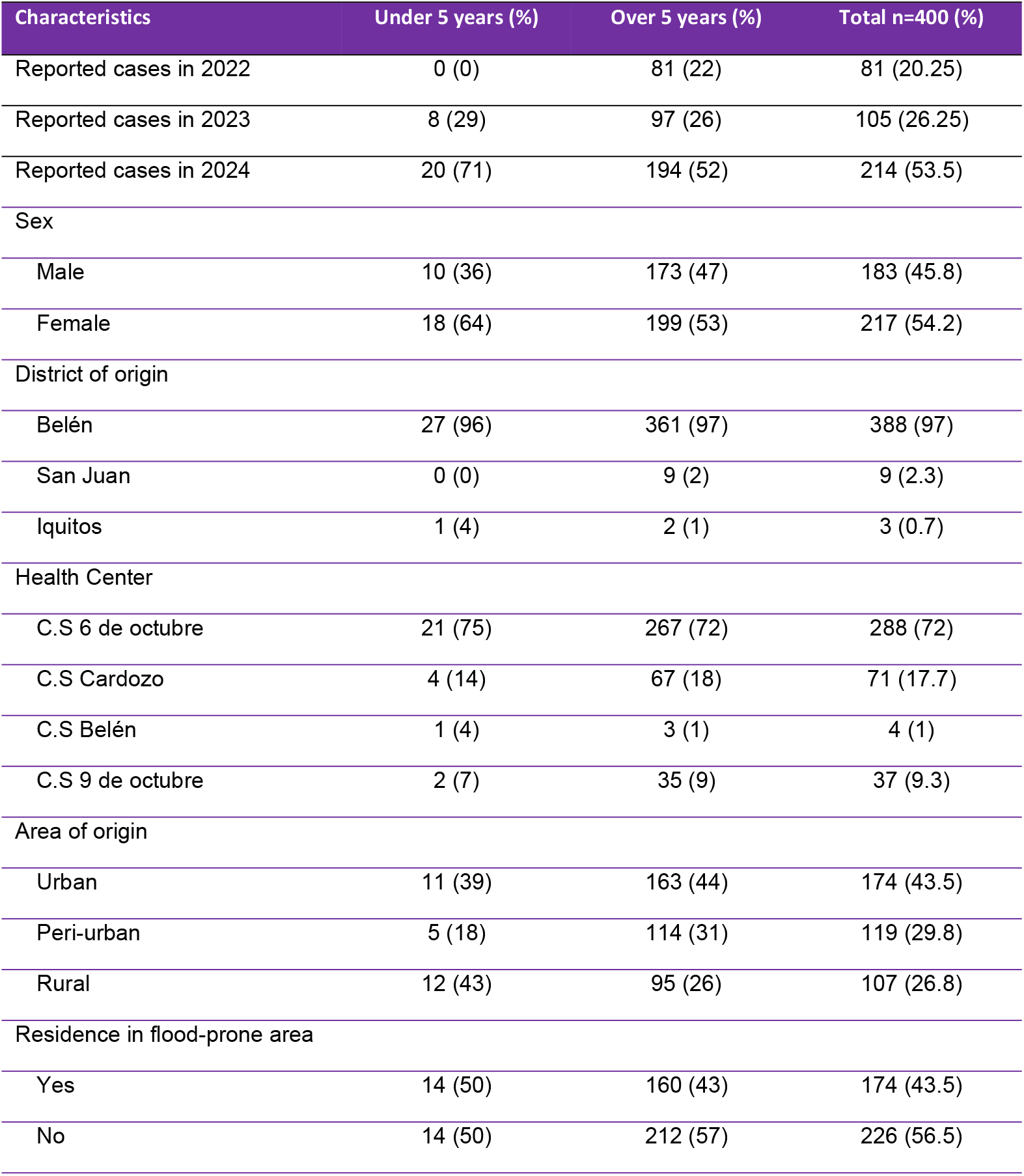
Epidemiological characteristics of patients diagnosed with Leptospirosis in Health Centers of the Peruvian Amazon, 2022–2024.

**Figure 1.**
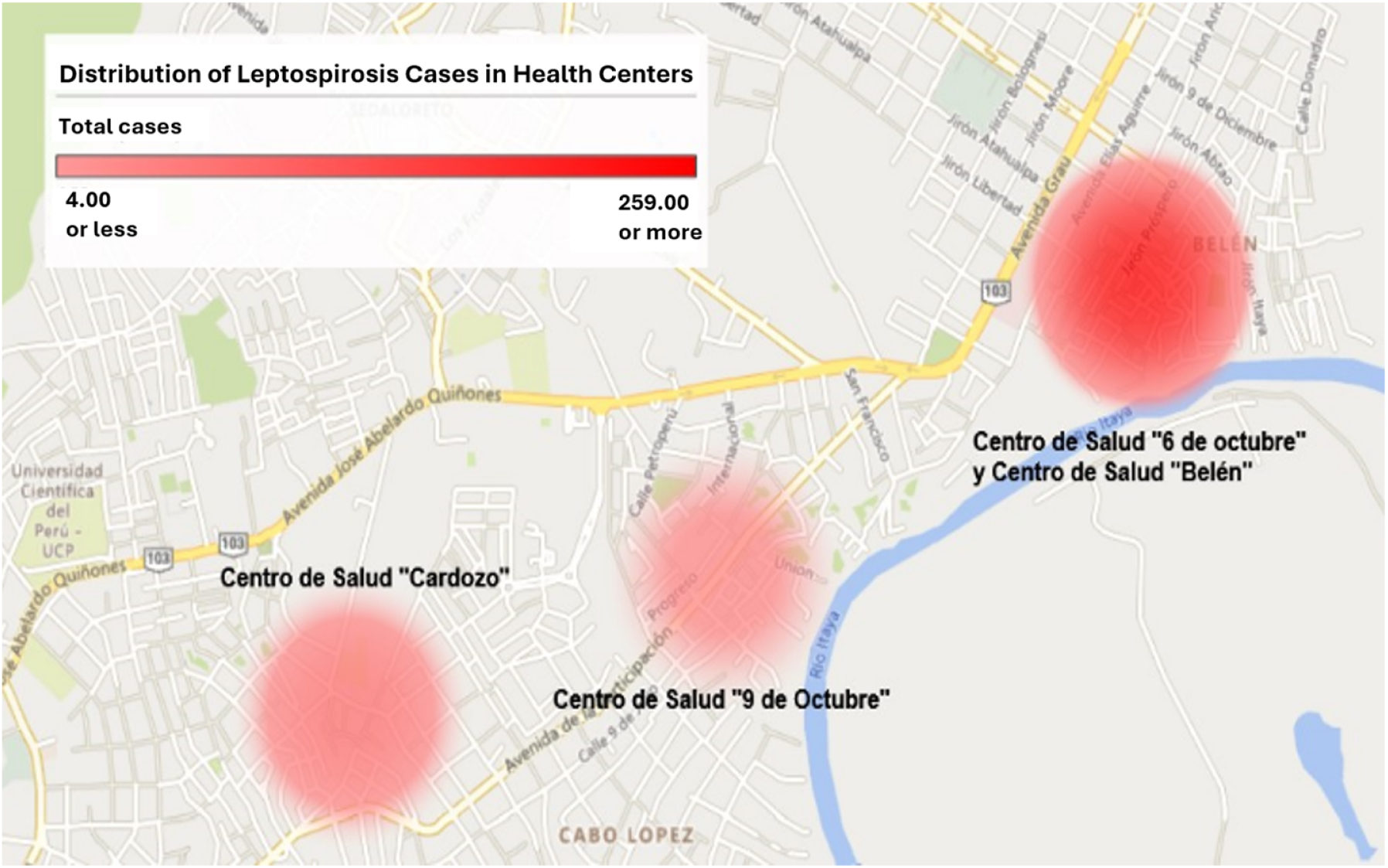
Distribution of Leptospirosis cases in Health Centers of the Peruvian Amazon, 2022–2024.

**Figure 2.**
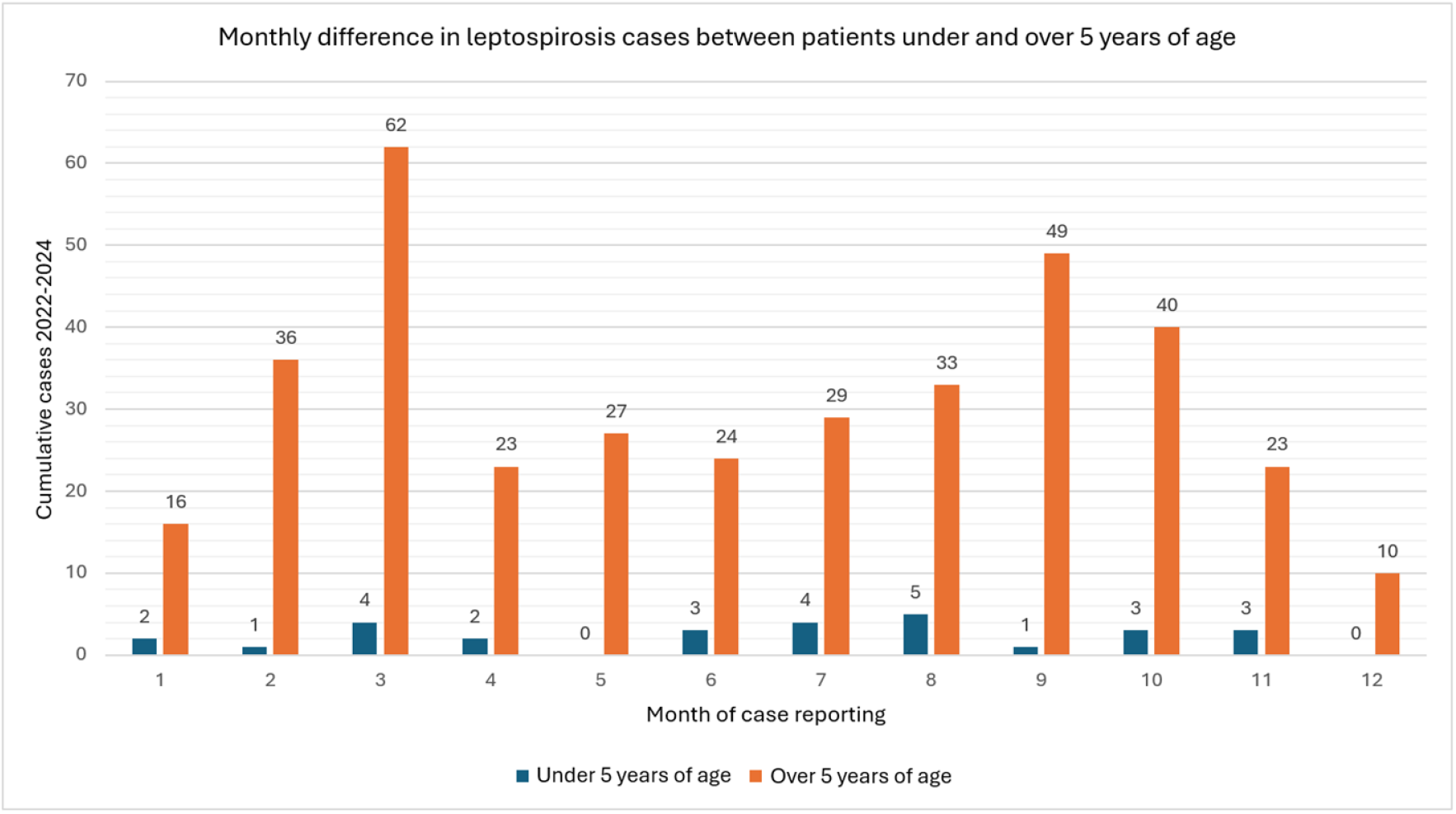
Monthly prevalence of leptospirosis cases in patients under and over 5 years of age in Health Centers of the Peruvian Amazon, 2022–2024.

Regarding the area of origin, 12 children under 5 years of age (43%) came from rural areas, followed by 11 (39%) from urban areas and 5 (18%) from peri-urban areas. In contrast, among individuals over 5 years old, urban origin predominated with 163 cases (44%), followed by 114 (31%) from peri-urban areas and 95 (26%) from rural areas. Fifty percent of the children under 5 resided in flood-prone areas, as did 43% of those over 5. The most affected sex was female in both groups, with 18 cases (64%) in children under 5 and 199 cases (53%) in those over 5. (Table 1)

The average time from the onset of illness to the date of consultation was between 3 to 6 days in 183 cases (45.8%). It was observed that the majority of cases in children under 5 years of age (46%) sought care within the first 3 days, while the older population (46%) was attended between days 3 to 6. In all cases, regardless of age group, the onset of illness was insidious and had a progressive course. (Table 2)

**Table 2.** Clinical characteristics of patients diagnosed with Leptospirosis in Health Centers of the Peruvian Amazon, 2022–2024.

| Characteristics | Under 5 years (%) | Over 5 years (%) | Total n=400 (%) |
| --- | --- | --- | --- |
| Duration of illness |  |  |  |
| 0 to 3 days | 13 (46) | 97 (26) | 110 (27.5) |
| 3 to 6 days | 12 (43) | 171 (46) | 183 (45.8) |
| More than 6 days | 3 (11) | 104 (28) | 107 (26.8) |
| Type of onset |  |  |  |
| Insidious | 28 (100) | 372 (100) | 400 (100) |
| Abrupt | 0 (0) | 0 (0) | 0 (0) |
| Course of illness |  |  |  |
| Intermittent | 0 (0) | 0 (0) | 0 (0) |
| Stationary | 0 (0) | 0 (0) | 0 (0) |
| Progressive | 28 (100) | 372 (100) | 400 (100) |
| Discharge status |  |  |  |
| Alive | 28 (100) | 372 (100) | 400 (100) |
| Deceased | 0 | 0 | 0 (0) |

Regarding symptoms, when comparing age groups, it was observed that the most nonspecific symptoms were more frequent in children under 5 years of age, with malaise (78.6%) and fever (71.4%) being the most common. In contrast, symptoms such as headache (66.4%), chills (35.8%), nausea/vomiting (23.1%), myalgia (16.4%), and musculoskeletal pain (10.2%) were more frequent in individuals over 5 years of age (Table 3).

**Table 3.**
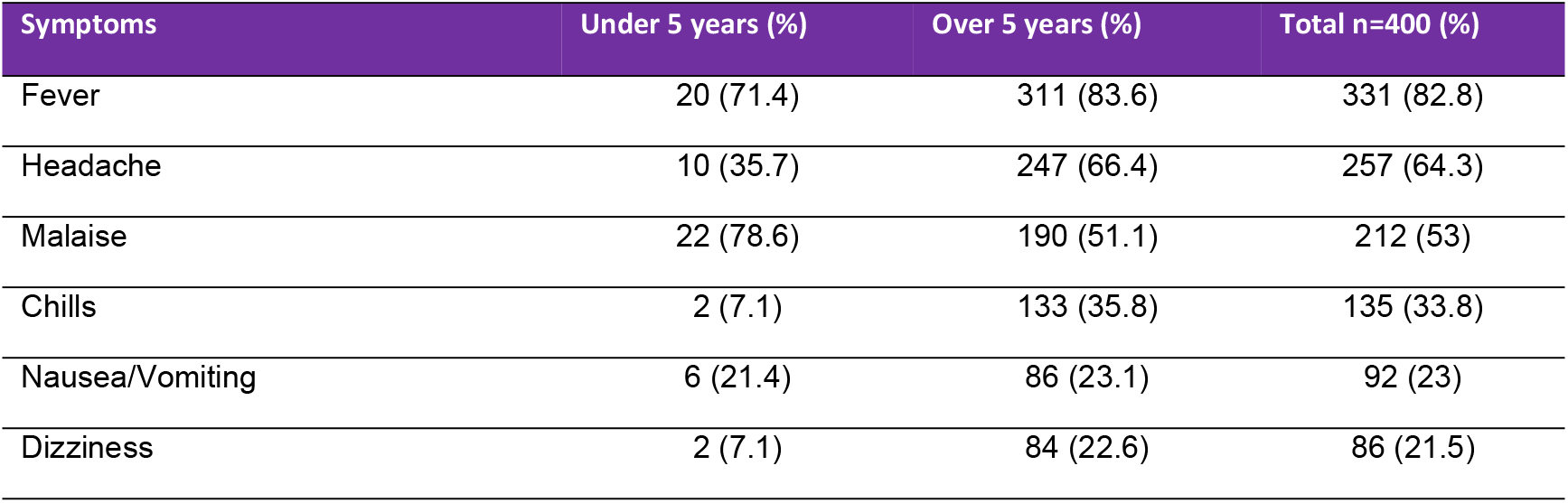
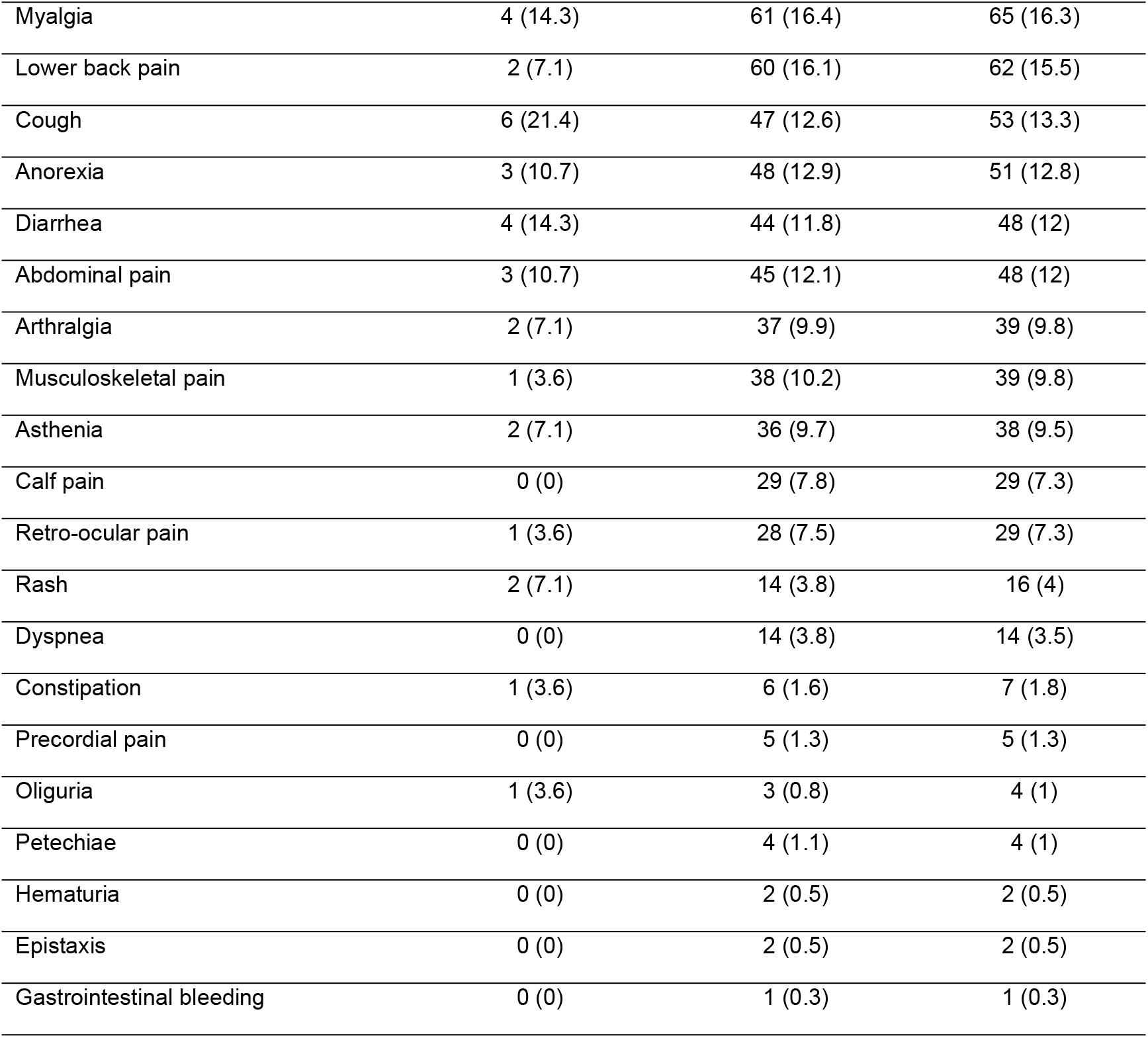
Frequency of symptoms presented by patients diagnosed with Leptospirosis in Health Centers of the Peruvian Amazon, 2022–2024.

| Symptoms | Under 5 years (%) | Over 5 years (%) | Total n=400 (%) |
| --- | --- | --- | --- |
| Fever | 20 (71.4) | 311 (83.6) | 331 (82.8) |
| Headache | 10 (35.7) | 247 (66.4) | 257 (64.3) |
| Malaise | 22 (78.6) | 190 (51.1) | 212 (53) |
| Chills | 2 (7.1) | 133 (35.8) | 135 (33.8) |
| Nausea/Vomiting | 6 (21.4) | 86 (23.1) | 92 (23) |
| Dizziness | 2 (7.1) | 84 (22.6) | 86 (21.5) |
| Myalgia | 4 (14.3) | 61 (16.4) | 65 (16.3) |
| Lower back pain | 2 (7.1) | 60 (16.1) | 62 (15.5) |
| Cough | 6 (21.4) | 47 (12.6) | 53 (13.3) |
| Anorexia | 3 (10.7) | 48 (12.9) | 51 (12.8) |
| Diarrhea | 4 (14.3) | 44 (11.8) | 48 (12) |
| Abdominal pain | 3 (10.7) | 45 (12.1) | 48 (12) |
| Arthralgia | 2 (7.1) | 37 (9.9) | 39 (9.8) |
| Musculoskeletal pain | 1 (3.6) | 38 (10.2) | 39 (9.8) |
| Asthenia | 2 (7.1) | 36 (9.7) | 38 (9.5) |
| Calf pain | 0 (0) | 29 (7.8) | 29 (7.3) |
| Retro-ocular pain | 1 (3.6) | 28 (7.5) | 29 (7.3) |
| Rash | 2 (7.1) | 14 (3.8) | 16 (4) |
| Dyspnea | 0 (0) | 14 (3.8) | 14 (3.5) |
| Constipation | 1 (3.6) | 6 (1.6) | 7 (1.8) |
| Precordial pain | 0 (0) | 5 (1.3) | 5 (1.3) |
| Oliguria | 1 (3.6) | 3 (0.8) | 4 (1) |
| Petechiae | 0 (0) | 4 (1.1) | 4 (1) |
| Hematuria | 0 (0) | 2 (0.5) | 2 (0.5) |
| Epistaxis | 0 (0) | 2 (0.5) | 2 (0.5) |
| Gastrointestinal bleeding | 0 (0) | 1 (0.3) | 1 (0.3) |

When analyzing the distribution of species by age group using the MAT test, the most frequent serological variety in both groups was *Varillal*, present in 86% of children under 5 years of age and 85% of those over 5. Among children, the *Panama* serotype stood out (7%) compared to older individuals (1%), while *Hurstbridge* was detected only in patients over 5 years old. In 21% of cases, antibodies were detected that reacted to more than two serological varieties with titers equal to or greater than 1:800, which could be due to cross-reactions between related serovars or multiple exposures. However, in 79% of the positive cases, a single predominant serological variety was identified, suggesting that most patients had an acute infection caused by a specific serovar. (Table 4)

**Table 4.** Types of species in patients diagnosed with Leptospirosis in Health Centers of the Peruvian Amazon.

| Characteristics | Under 5 years (%) | Over 5 years (%) | Total n=400 (%) |
| --- | --- | --- | --- |
| Species |  |  |  |
| *Varillal* | 24 (86) | 316 (85) | 340 (66.9) |
| *Hurstbridge* | 0 (0) | 47 (13) | 47 (9.3) |
| *Icterohemorrhagiae* | 2 (7) | 41 (11) | 43 (8.5) |
| *Bratislava* | 3 (11) | 28 (8) | 31 (6.1) |
| *Copenhag* | 1 (4) | 12 (3) | 13 (2.6) |
| *Australis* | 1 (4) | 7 (2) | 8 (1.6) |
| *Panama* | 2 (7) | 4 (1) | 7 (1.4) |
| *Autumnalis* | 0 (0) | 5 (1) | 5 (1) |
| *Canicola* | 0 (0) | 4 (1) | 4 (0.8) |
| *Bataviae* | 0 (0) | 3 (0.8) | 3 (0.6) |
| *Cynopteri* | 1 (4) | 2 (0.5) | 3 (0.6) |
| *Coxi* | 0 (0) | 1 (0.2) | 1 (0.2) |
| *Javanic* | 0 (0) | 1 (0.2) | 1 (0.2) |
| *Hardjo* | 0 (0) | 1 (0.2) | 1 (0.2) |
| *Djasiman* | 0 (0) | 1 (0.2) | 1 (0.2) |
| Patients with a single species | 23 (82) | 294 (79) | 317 (79.25) |
| Patients with two species | 4 (14) | 54 (15) | 58 (14.5) |
| Patients with three species | 1 (4) | 23 (6) | 24 (6) |
| Patients with four species | 0 (0) | 1 (0) | 1 (0.25) |

Regarding treatment, it was observed that the most commonly used antipyretic was Paracetamol, administered in 303 cases (75.8%), used in 61% of children under 5 years and in 77% of those over 5 years. Among antibiotics, amoxicillin was the most frequently used in children under 5 (71%), while doxycycline predominated in the older group (60%). Antibiotics such as ciprofloxacin, ceftriaxone, and ampicillin were used exclusively in patients over 5 years of age, whereas erythromycin was used more frequently in children under 5 (11%) than in older individuals (4%). Intravenous administration of 0.9% sodium chloride was provided in 11% of children under 5 and in 13% of those over 5. (Table 5)

**Table 5.** Medications used in patients diagnosed with Leptospirosis in Health Centers of the Peruvian Amazon, 2022–2024.

| Medications | Under 5 years (%) | Over 5 years (%) | Total n=400 (%) |
| --- | --- | --- | --- |
| Antipyretics |  |  |  |
| Paracetamol | 17 (61) | 286 (77) | 303 (75.8) |
| Metamizole | 4 (14) | 47 (13) | 51 (13) |
| Antibiotics |  |  |  |
| Doxycycline | 5 (18) | 222 (60) | 227 (56.8) |
| Amoxicillin | 20 (71) | 95 (26) | 115 (28.8) |
| Ciprofloxacin | 0 (0) | 35 (9) | 35 (8.8) |
| Erythromycin | 3 (11) | 14 (4) | 17 (4.3) |
| Ceftriaxone | 0 (0) | 10 (3) | 10 (2.5) |
| Ampicillin | 0 (0) | 2 (1) | 2 (0.5) |
| IV hydration |  |  |  |
| 0.9% sodium chloride | 3 (11) | 49 (13) | 52 (12.8) |
| Others |  |  |  |
| Dimenhydrinate | 1 (4) | 14 (4) | 15 (3.8) |

In addition to the descriptive analysis, a bivariate analysis was conducted using Pearson’s Chi-square test to identify clinical and epidemiological variables significantly associated with the age group of under vs. over 5 years (Table 6).

**Table 6.**
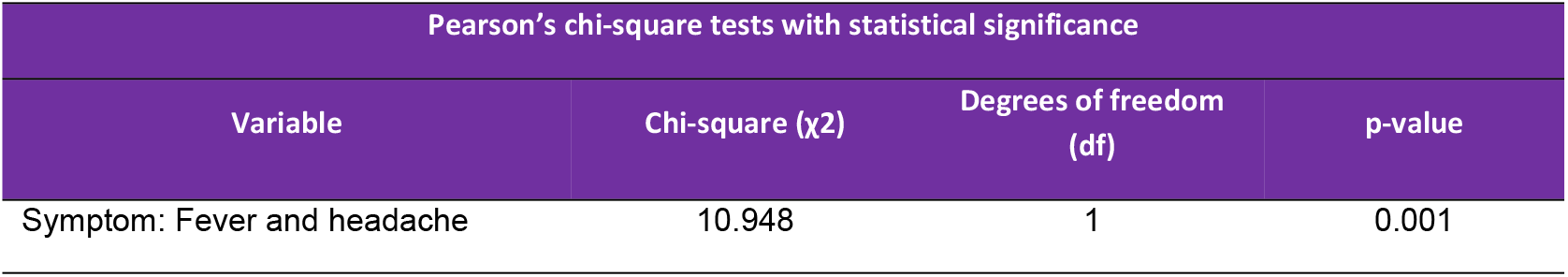
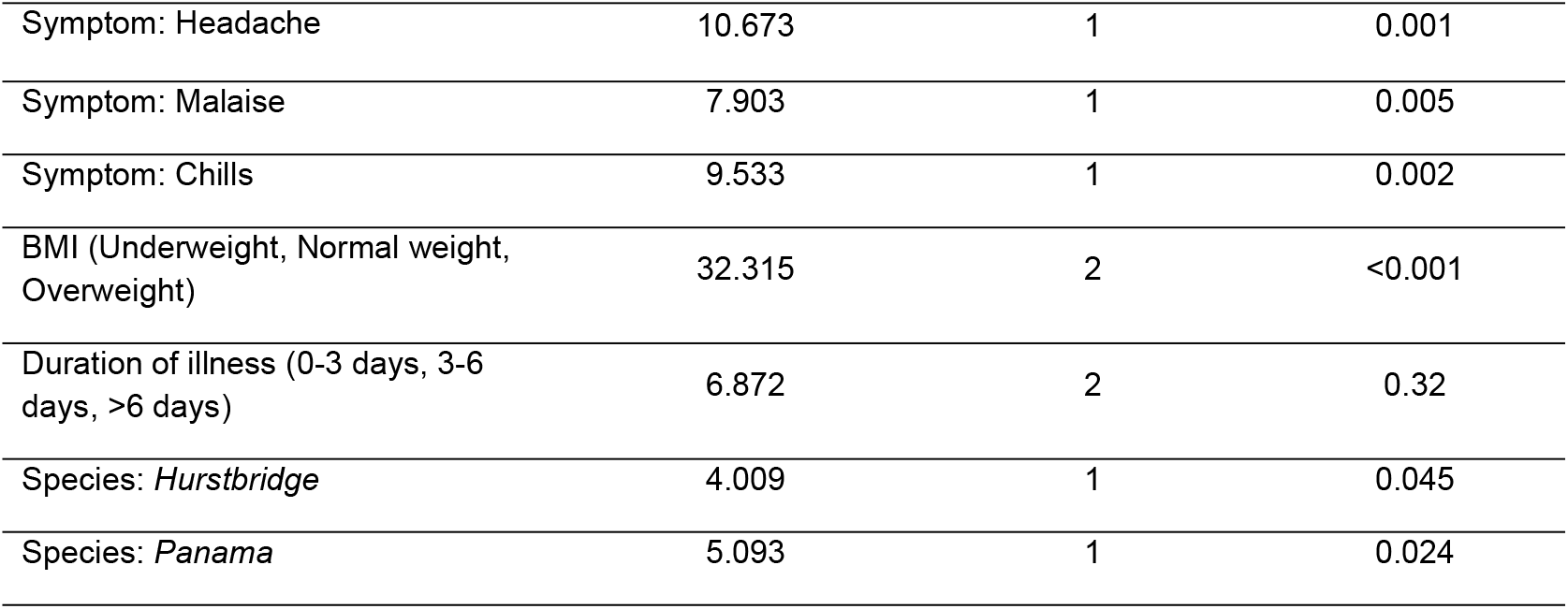
Bivariate analysis of characteristics according to age group: under 5 years vs over 5 years.

A statistically significant association was found for the combined symptom of fever and headache, being more frequent in the group over 5 years of age (χ^2^ = 10.948; p = 0.001), as well as for isolated headache (χ^2^ = 10.673; p = 0.001) and chills (χ^2^ = 9.533; p = 0.002). On the other hand, a significant association was identified with the symptom of malaise, which was more prevalent in the group under 5 years of age (χ^2^ = 7.903; p = 0.005).

Regarding nutritional status, a highly significant association was found between BMI (underweight, normal weight, and overweight) and age group, with a higher proportion of underweight observed in children under 5 years of age (χ^2^ = 32.315; p = <0.001). The categorized duration of illness (0–3 days, 3–6 days, >6 days) did not show a statistically significant association (χ^2^ = 6.872; p = 0.32). With respect to *Leptospira* species, a statistically significant association was observed with the *Hurstbridge* species, which was found exclusively in individuals over 5 years of age (χ^2^ = 4.009; p = 0.045), while the *Panama* species was more frequent in the group under 5 years of age (χ^2^ = 5.093; p = 0.024).

In the multivariate analysis using binary logistic regression to identify factors associated with the under-5 age group, two variables showed statistical significance. Being underweight according to BMI was significantly associated with a higher probability of belonging to the group under 5 years of age (OR = 0.051; p < 0.001), indicating an inverse association: underweight patients had a 94.9% reduction in the probability of belonging to the older group, meaning a strong association with being under 5 years of age. Likewise, a longer duration of illness was associated with an increased probability of belonging to the under-5 group (OR = 1.366; p = 0.001), suggesting that for each additional day of symptoms, the likelihood of being in the under-5 group increased by approximately 36.6%. (Table 7)

**Table 7.** Multivariate Analysis Using Binary Logistic Regression to Identify Factors Associated with the Under-5 Age Group.

| Binary Logistic Regression: Children Under 5 Years vs. Over 5 Years |  |  |  |  |  |  |
| --- | --- | --- | --- | --- | --- | --- |
| Variable | Regression coefficient (B) | Standard error | Wald statistic | df | p-value | Odds ratio (Exp (B)) |
| BMI: underweight | -2.969 | 0.780 | 14.473 | 1 | <0.001 | 0.051 |
| Duration of illness (quantitative) | 0.312 | 0.98 | 10.160 | 1 | 0.001 | 1.366 |

## Discussion

The present study provides relevant data on the epidemiological and clinical behavior of leptospirosis in the Amazon region of Peru, with emphasis on the differences between patients under and over 5 years of age.

The present study reveals a defined seasonal pattern in the population over 5 years of age, which coincides with the climatic cycle of the district of Belén, characterized by periods of river rise and intense rainfall that create favorable conditions for the proliferation of *Leptospira*. This temporal association reinforces the role of the environment as a determining factor in transmission. In contrast, children under 5 years of age did not exhibit a defined seasonal pattern, which could be explained by lower direct exposure to contaminated water during flooding or by reduced behavioral susceptibility, as this age group generally has limited mobility or remains in more controlled environments.

Factors such as the presence of nonspecific symptoms, underweight BMI, and duration of illness of 0–3 days were identified as significant findings regarding the epidemiological and clinical behavior of leptospirosis in pediatric patients under 5 years of age compared to those over 5 years, treated at primary health care centers in the district of Belén, Loreto region, during the 2022–2024 period. This population, historically underrepresented in leptospirosis studies, presents distinct clinical and social characteristics that should be considered to improve early diagnosis and the comprehensive management of the disease.

One of the most relevant findings was the high frequency of nonspecific symptoms in children under 5 years of age, particularly malaise, reported in 78.6% of the cases in this age group. This contrasts with the population over 5 years, in which more classic symptoms such as fever, headache, and chills predominated. This symptomatic difference has also been documented in studies conducted in similar settings, where it is noted that young children tend to present with atypical or less specific clinical forms, which may hinder timely diagnosis if a high level of clinical suspicion is not maintained in endemic areas [6,14].

The clinical profile of children affected by leptospirosis in this study is characterized by the presentation of vague and non-specific symptoms, which may contribute to underreporting of cases in the pediatric group. This highlights the importance of training healthcare personnel in the recognition of subtle clinical presentations in vulnerable populations and in areas with high endemicity [15].

Another notable aspect was the high frequency of underweight status according to BMI in children under 5 years of age, which showed a statistically significant association with age (p < 0.001). This finding suggests a double burden of vulnerability: on one hand, environmental exposure to a zoonotic pathogen such as *Leptospira*, and on the other, the presence of chronic or acute malnutrition as a reflection of precarious socioeconomic conditions. Malnutrition compromises the immune response and could influence the less florid clinical presentation observed in this group, in addition to affecting the course and recovery of the cases [16,17].

Likewise, it was identified that the duration of illness prior to consultation was shorter in children under 5 years of age, with 46% of them seeking care at a health facility within the first three days of symptom onset. This early healthcare-seeking behavior may be explained by the concern of caregivers in response to signs of illness in infants and preschoolers, who are unable to communicate complex symptoms such as headache or musculoskeletal pain, once again reinforcing the predominance of nonspecific symptoms such as fever and fatigue [7,14,18].

From a microbiological perspective, the predominant serovar in both age groups was *Varillal*. However, the notable finding was the *Panama* serovar, which was significantly more frequent in children under 5 years of age (7% vs. 1%; p = 0.024). This may have implications in terms of virulence, transmission patterns, and immunological susceptibility in early childhood. Although additional studies are needed to confirm this finding, the identification of specific serovars in certain age groups could be useful for designing preventive strategies and developing future vaccines [19].

Regarding treatment, antibiotic selection also reflected age-related differences. In younger children, amoxicillin was the preferred antibiotic (71%), in accordance with treatment guidelines recommended for pediatric populations [17]. In contrast, doxycycline predominated in patients over 5 years of age (60%). These patterns reflect appropriate clinical judgment in antibiotic prescription according to age group, which is crucial to prevent renal, hepatic, or pulmonary complications associated with severe forms of the disease [7,20].

Finally, it is important to consider that all cases included in this study correspond to mild forms of leptospirosis, with no reports of hospitalization or death. Although this limitation reduces the ability to generalize findings to severe cases, it provides a valuable overview of the outpatient profile of the disease in a highly vulnerable Amazonian district such as Belén.

The clinical profile of children under 5 years of age with leptospirosis in this series is characterized by: predominance of female sex, rural or peri-urban origin, residence in flood-prone areas, nonspecific symptoms (malaise and fever), low BMI, early consultation (0–3 days), and predominance of the Varillal serovar followed by Panama. These findings justify the need to strengthen clinical and epidemiological surveillance in this age group, promoting differentiated strategies for prevention, early diagnosis, and appropriate treatment in endemic areas.

### Limitations

No deaths were reported among the analyzed cases, as the data correspond to mild forms of leptospirosis treated at primary-level health facilities. Cases with severe clinical manifestations or complications are referred from primary care to higher-level centers. The exclusive inclusion of mild cases limits the ability to generalize the findings to patients with severe forms of the disease, who may require specialized management and present adverse outcomes.

## Data Availability

All data underlying the findings of this study are fully available without restriction and are provided within the manuscript and its Supporting Information files.

